# Uterine Fibroids and Hypertensive Disorders in Pregnancy: A Systematic Review and Meta-Analysis

**DOI:** 10.1101/2024.03.05.24303824

**Authors:** Susan Nasab, Ethan K Gough, Elisabeth Nylander, Mostafa Borahay, James Segars, Valerie Baker, Xiaobin Wang, Katherine Cameron

## Abstract

**Capsule:** In this study the presence of uterine fibroids was significantly associated with an increased risk of development of hypertensive disorders of pregnancy even when accounting for age and BMI in meta-regression. This finding has potential implications for risk stratification and monitoring for hypertension during pregnancy in this population.

**Objective:** To examine the association between uterine fibroids and the development of hypertensive disorders in pregnancy.

**Data sources:** Cochrane, Embase, PubMed, MEDLINE, Scopus, and Web of Science databases were searched from inception through April 2023.

**Study Selection and Synthesis:** Cohort, case-control, or case series studies including uterine fibroid status and hypertensive disorders of pregnancy status were included. The comparison group was pregnant women without uterine fibroids. Inverse-variance weighted random effects models were used to pool RR and OR estimates separately. Age and BMI were explored as potential sources of heterogeneity using inverse-variance weighted meta-regression.

**Main Outcomes:** Hypertensive disorders of pregnancy (HDP) defined as gestational hypertension, pre-eclampsia, eclampsia, superimposed preeclampsia, or hemolysis, elevated liver enzymes, and low platelets (HELLP) syndrome.

**Results:** A total of 17 studies were included (Total N=1,374,395 participants, N=64,968 with uterine fibroids). Thirteen studies were retrospective cohorts and four were case-control studies. Women with uterine fibroids had a significantly higher risk of hypertensive disorders in pregnancy compared to women without uterine fibroids with RR 1.74 (95% CI 1.33-2.27, p<0.01), and OR 2.87 (95% CI 1.38-5.97, p<0.01), in cohort studies and case-control studies, respectively. In meta-regression analyses, age did not significantly change the positive association between uterine fibroids and hypertensive disorders in pregnancy.

**Conclusion:** Uterine fibroids were associated with an increased risk of hypertensive disorders of pregnancy when all available literature was synthesized, including when shared risk factors are examined in meta-regression analyses.

**Relevance:** If confirmed in future studies, investigations into the mechanisms of this association are needed as this finding potentially has implications for risk stratification and monitoring for hypertensive disorders of pregnancy in this population.

**Trial Registration:** PROSPERO, ID # 331528

## INTRODUCTION

Uterine leiomyomas (fibroids) are the most common solid symptomatic neoplasm in women, estimated to occur in up to 70% of women by the time of menopause (1). Uterine fibroids are associated with morbidities including bulk symptoms such as pelvic pressure, urinary frequency, and constipation, as well as abnormal uterine bleeding, anemia, and infertility (2). In addition, uterine fibroids have been associated with poor obstetrical outcomes such as spontaneous abortion, fetal malpresentation, preterm labor, postpartum hemorrhage, and cesarean section (3–6).

Increasing data supports an association between uterine fibroids and chronic hypertension (7). However, it is unclear whether this association is due to common risk factors or underlying mechanisms - both conditions share commonalities including alterations of smooth muscle cell functioning, hypoxia, and angiogenesis (8–11). In addition, there is conflicting literature regarding whether there is an association between uterine fibroids and the development of hypertensive disorders in pregnancy. Fibroids are more prevalent and occur at younger ages in black women, who also have a higher rate of maternal morbidity and mortality. Since pregnancy may be viewed as a “cardiac stress test”, with the development of preeclampsia in pregnancy linked to the development of hypertension long-term, understanding the association between uterine fibroids and hypertensive disorders in pregnancy has important implications for both offspring and maternal health. Therefore, we sought to perform a systematic review and meta-analysis of existing studies to evaluate the association between uterine fibroids and hypertensive disorders of pregnancy.

## MATERIALS AND METHODS

This systematic review and meta-analysis followed the Preferred Reporting Items for Systematic Reviews and Meta-Analyses (PRISMA) guidelines. The study protocol was registered in the international prospective register of systematic reviews (PROSPERO, registration ID: 331528 on 1/10/2022). Funding supports for this study was WRHR NIH NICHD Award # K12 HD103036, PI Andrew Satin, RD James Segars.

### Search Strategy

Electronic databases were searched using Cochrane, Embase, PubMed, MEDLINE, Scopus, and Web of Science from inception to April 2023. An advanced search was done with the MeSH terms listed in Supplement 1.

### Inclusion and Exclusion Criteria

Criteria for inclusion in the study were established a priori. All studies available in English language text including retrospective or prospective cohort studies, case-control, or case series that included information regarding uterine fibroid status prior to pregnancy and the outcome of hypertensive disorders of pregnancy and contained a comparison group were included. Case reports, reviews, case studies without comparison groups, and studies of multifetal gestations were excluded.

### Outcome Measure

Hypertensive disorders of pregnancy were the primary study outcome. Hypertensive disorders of pregnancy included gestational hypertension, pre-eclampsia, eclampsia, superimposed preeclampsia, or HELLP syndrome (Hemolysis, Elevated Liver enzymes, and Low Platelets), and the definition utilized by each individual study is described in Supplement 2. HDP rates were extracted from each study for the fibroid and unaffected groups.

### Data Extraction

Two independent reviewers (S.N and K.C) reviewed all studies identified from a primary search using the predefined inclusion and exclusion criteria. After the removal of duplicates, this included title screening with abstract and tables review, followed by a full-text review of relevant articles. Search outcomes were carefully recorded using a reference manager (Covidence). Discrepancies were resolved by consensus.

### Quality Assessment

The risk of bias in the included studies was assessed independently by two independent reviewers (S.N and K.C). The Newcastle-Ottawa Scale (NOS) was used to assess the quality of included studies according to the study type. The NOS comprises “participant selection,” “comparability of study groups,” and “assessment of outcome or exposure.” A score > or equal to 7 is considered high quality.

### Statistical Analysis

We estimated relative risks (RR) from cohort studies and odds ratios (OR) from case-control studies. We used inverse-variance weighted random effects models to pool RR and OR estimates separately and used forest plots for visualization. We assessed statistical heterogeneity using the I^2^ statistic. We explored mean age and mean BMI as potential sources of heterogeneity using inverse-variance weighted meta-regression.

Mean age was estimated from reported age group frequencies for two studies (12, 13), assuming lower and upper limits of 18 and 45, respectively. Mean BMI was estimated from reported BMI category frequencies for one study (4), assuming the same lower and upper limits. We assessed publication bias using Egger’s test and funnel plots. Statistical significance was evaluated at α<0.05. Further sensitivity analyses were performed to determine the robustness of pooled effect estimates to the exclusion of individual studies and with low quality studies censored. All analyses were conducted using the Metafor package, in R version 4.2.2.(14)

## RESULTS

A total of 6458 studies were identified for screening. Of those, 2958 were screened after removing the duplicates. One hundred forty-eight full-text studies were assessed for eligibility and reviewed in detail. Seventeen studies met the inclusion criteria and were included in the final meta-analysis. (Figure 1)

**Figure 1:**
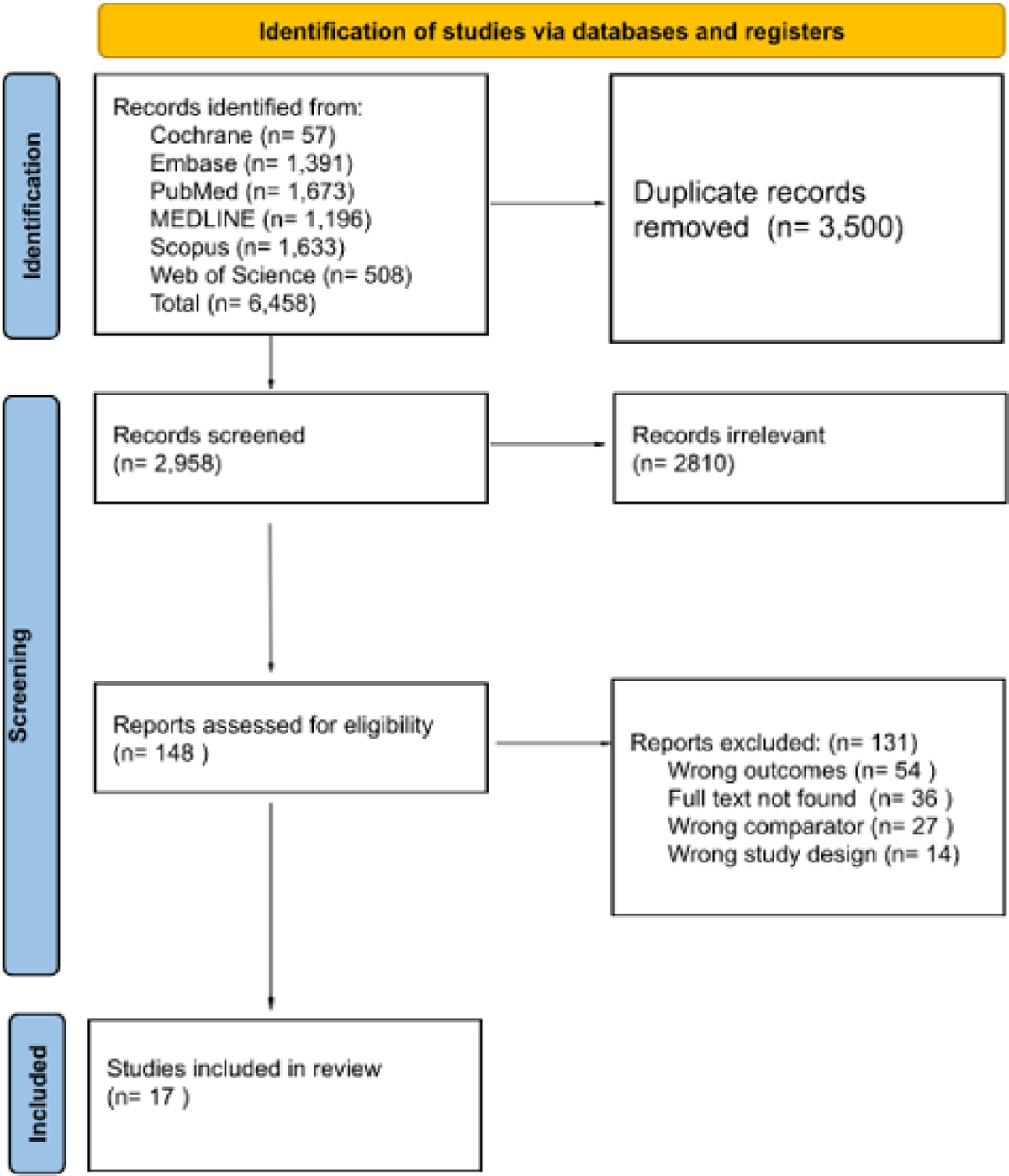
PRISMA Flow Diagram

### Study Characteristics

A total of 1,374,395 women were included in the final meta-analysis from studies published from 1998-2022. Of those 1,374,395 women, 64,968 were diagnosed with uterine fibroids. Six studies were performed in China (13, 15–19), five studies in the United States (4, 6, 12, 20, 21), 2 in Israel (22, 23), 1 in Italy (24), 1 in France (25), 1 in Korea (26), and 1 in Australia (27). Thirteen studies were designed as a retrospective cohort (4, 6, 12, 15, 17, 19–26) and 4 as case-control (13, 16, 18, 27). One study included an intervention arm resulting in analysis by three groups (no uterine fibroid, fibroid with myomectomy prior to pregnancy, and fibroid without myomectomy) (26). Myomectomy was considered an intervention. We therefore performed all analyses twice — with or without the inclusion of participants in the intervention group in Lee et al (26) — to determine the robustness of our results. A summary of each study’s characteristics, including inclusion criteria and demographic characteristics, is presented in Table 1.

**Table 1.** Clinical Characteristics of the Studies Included for Meta-Analysis.

| Author | Year | Country | Study design | Inclusion Criteria | Total | Fibroid Diagnosis | Age<br>Fibroid<br>No Fibroid<br>P Value | BMI/Weight/<br>Obesity<br>Fibroid<br>No Fibroid<br>P Value | AA race<br>Fibroid<br>No Fibroid<br>P Value | Newcastle<br>Ottawa Scale<br>(NOS) |
| --- | --- | --- | --- | --- | --- | --- | --- | --- | --- | --- |
| Biderman<br>-Madar | 2005 | Israel | Retrospective | Population based study,<br>women conceived after<br>fertility treatment, singleton<br>gestation, delivered in Soroka<br>University Medical Center | 1995 | Medical<br>Records | Mean+/-SD<br>34.5+/-5.5<br>29.7+/-5.2<br><0.001 | NR | NR | 7 |
| Chen | 2021 | China | Retrospective | Women who received prenatal<br>care in the first affiliated<br>hospital of Shantou University<br>Medical College, 20-45 years<br>old, women who had<br>confirmation of pregnancy via<br>ultrasound before 20th week<br>of gestation | 2277 | Ultrasound | Mean+/-SD<br>32.3 +/-5.0<br>30.3+/-4.4<br><0.001 | Mean+/-SD<br>21.5+/-3.1<br>20.4+/-3.0<br><0.001 | NR | 9 |
| Conti | 2013 | Italy | Retrospective | Multicenter study, Caucasian<br>primipara women, > 30 years<br>old, delivered in Siena and<br>Florence, 2011 | 450 | Survey | N>35 years<sup>a</sup><br>105<br>60<br>0.0004 | Mean<br>weight(kg) +/-<br>SD<br>61+/-10<br>63+/-14<br>NS | NR | 5 |
| Coronado | 1998 | United States | Retrospective | Population based study, data<br>from Washington states birth<br>certificates linked to hospital<br>discharge records was used,<br>singleton live births, 1987-1993 | 6308 | ICD9 | NR | NR | N (%)<br>202 (9.8)<br>159 (3.7)<br>NR | 9 |
| Farland | 2022 | United States | Retrospective | Data from SART CORS, PELL, APCD were linked for the analysis., ART cycles in the state of Massachusetts 2004-2017 were linked to delivery and hospital discharge records* | 91825 | ICD9, ICD10 | Mean+/-SD<br>35.78+/-4.17<br>33.00+/-4.03<br>NR | NR | N (%)<br>429 (10.2)<br>2154 (2.9)<br>NR | 9 |
| Girault | 2018 | France | Retrospective | Women delivered a singleton fetus > 22 weeks of gestation, in a tertiary university hospital, 2011-2015 | 19866 | Ultrasound | Mean+/-SD<br>36.1+/-5.0<br>32.0+/-5.4<br><0.001 | Mean+/-SD<br>25.4+/-5.4<br>22.8+/-4.6<br><0.001 | NR | 7 |
| Harlev | 2019 | Israel | Retrospective | Population based study, singleton delivery, single regional tertiary medical center, 1991-2014 | 242445 | Medical Records | Mean+/-SD<br>34.22+/-5.5<br>28.13+/-5.8<br>0.003 | Obese<sup>b</sup> N (%)<br>33 (2.7)<br>2424 (1.0)<br><0.001 | NR | 7 |
| Knight | 2017 | United States | Retrospective | Women with singleton pregnancies, undergoing 2nd trimester fetal anatomy ultrasound at Riverside Methodist Hospital, Ohio, 2007-2012 | 282 | ICD9 | Mean+/-SD<br>34.2+/-4.4<br>31.5+/-5.2<br><0.001 | Mean+/-SD<br>29.4+/-8.0<br>27.3+/-7.1<br>0.023 | N (%)<br>47 (33)<br>44 (31)<br>0.95 | 7 |
| Lee | 2020 | Korea | Retrospective | Population based study, national health insurance (KNHI)** database was used, 2004-2015 | 788967 | ICD10 | NR | NR | NR | 9 |
| Roberts | 1999 | United States | Retrospective | Women with initial ultrasound study before 20 weeks of gestation, data from antenatal diagnostic unit (ADU) was used, delivery at the university | 153 | Ultrasound | Mean+/-SD<br>31.1 +/-5.7<br>23.6+/-6.3 | Mean weight (kg) +/-SD<br>89.1+/-23.0<br>82.9+/-23.6 | N (%)<br>50 (98)<br>75 (74) | 7 |
|  |  |  |  | of Mississippi medical center, 1994-1995 |  |  | 0.001 | NS | 0.001 |  |
| Stout | 2010 | United States | Retrospective | Women with singleton pregnancy, underwent routine 2nd trimester fetal anatomy ultrasound, single center, Washington University, 1990-2007 | 64047 | Ultrasound | Mean+/-SD<br>35.1+/-4.6<br>30+/-6.3<br><0.01 | Mean+/-SD<br>25.8+/-9.5<br>24.5+/-9.07<br><0.01 | N (%)<br>710 (34.5)<br>12583 (20.3)<br><0.01 | 9 |
| Wang | 2022 | China | Retrospective | Electronic medical records from singleton pregnant women diagnosed with adenomyosis and delivered at Peking University People's Hospital, 2015-2020. | 53 | Medical records | Mean+/-SD<br>35.4+/-4<br>34.8+/-3.8<br>0.551 | Mean+/-SD<br>24.5+/-3.6<br>23.2+/-3<br>0.184 | NR | 6 |
| Zhao | 2017 | China | Retrospective | Multicenter study ( 39 hospitals in 14 provinces in China), delivery in 2011 | 112403 | Ultrasound | Mean+/-SD<br>32+/-4.9<br>27.9+/-5.2<br><0.01 | Mean+/-SD<br>22.4+/-3.1<br>21.6+/-3.0<br><0.01 | NR | 9 |
| Chung | 2021 | Australia | Case-Control | Population-based study, Women born in 1921-26, 1946-51 and 1973-78, randomly selected from the national Medicare database, Australian citizens and permanent residents | 4473 | Survey | Mean+/-SD<br>24.7+/-1.4<br>24.6+/-1.4<br>0.1 | Obese<sup>b</sup> N (%)<br>74 (15.5)<br>283 (17.8)<br><0.01 | NR | 5 |
| Gong | 2022 | China | Case-Control | Electronic medical records from women who delivered at the People's Hospital of Xinjiang Uygur Autonomous | 699 | Ultrasound and self-report | Mean+/-SD<br>33+/-4.4 | Mean weight (kg) +/-SD<br>30.8+/-4.5 | NR | 9 |
|  |  |  |  | Region, Urumqi were used. Age at least 18 years old, singleton delivery with live newborn, available ultrasound results from early in pregnancy |  |  | 32.9+/-4.2<br>0.797 | 27.6+/-4<br><0.001 |  |  |
| Pan | 2019 | China | Case-Control | Discharge records from 2010-2015 of the third affiliated hospital of Guangzhou Medical University were used, patients screened with diagnosis of at least one of the following: Preeclampsia, HELLP syndrome, eclampsia or delivery | 6152 | Medical Records | <Age 30 years, N (%)<br>755 (46)<br>2657 (59)<br><0.01 | Obese<sup>b</sup> N (%)<br>273 (16.5)<br>254 (5.6)<br><0.001 | NR | 9 |
| Yi | 2017 | China | Case-Control | Singleton delivery at Wuhan medical and healthcare center, 2006-2015 | 32000 | Medical Records | Mean+/-SD<br>29+/-5.2<br>28+/-4.3<br>NR | Mean weight<br>83+/-3.7<br>71+/-3.6<br>NR | NR | 7 |

a: Number of participants. All included patients were above the age of 35 years old.
b: Number of obese patients, BMI>30
For case-control studies the variable is reported in case (those with hypertensive disorders of pregnancy) and control (those without hypertensive disorders of pregnancy) groups.
\*Society for Assisted Reproductive Technology Clinic Outcome Reporting System (SART CORS), Massachusetts Pregnancy to Early Life Longitudinal (PELL), Massachusetts all-payer claims Database (APCD), Assisted Reproductive Technology (ART)
\*\* The Korea National Health Insurance (KNHI)
NS: Not significant
NR: Not reported
HDP: Hypertensive disorders of pregnancy

### Meta-Analysis

Women with uterine fibroids had a significantly higher risk of hypertensive disorders in pregnancy compared to women without uterine fibroids (RR 1.74 [95% CI 1.33-2.27, p<0.01], OR 2.87 [95% CI 1.38-5.97, p<0.01]), in cohort studies and case-control studies, respectively (Figure 2). A similar statistically significant association was noted among cohort studies when the intervention arm of Lee et al (26) was excluded from the analysis (Supplement 3). In multivariable meta-regression models, age did not statistically significantly moderate the association between uterine fibroids and hypertensive disorders in pregnancy when controlling for BMI, but the impact of a uterine fibroid diagnosis was more significant in increasing the risk of HDP in women with lower compared to higher BMI when controlled for age (P-value 0.385, and 0.029 for age and BMI, respectively) (Supplement 4A, 4B). In other words, when controlling for age, the impact of the presence of uterine fibroids on the risk of HDP in women with larger BMI was only a small increase, whereas when controlled for age, the impact of the presence of uterine fibroids on the risk of HDP in women with smaller BMI was more pronounced. Finally, the above-mentioned findings did not change when the Lee et al (26) study was excluded from the analysis. (Supplement 5A, 5B)

**Figure 2.**
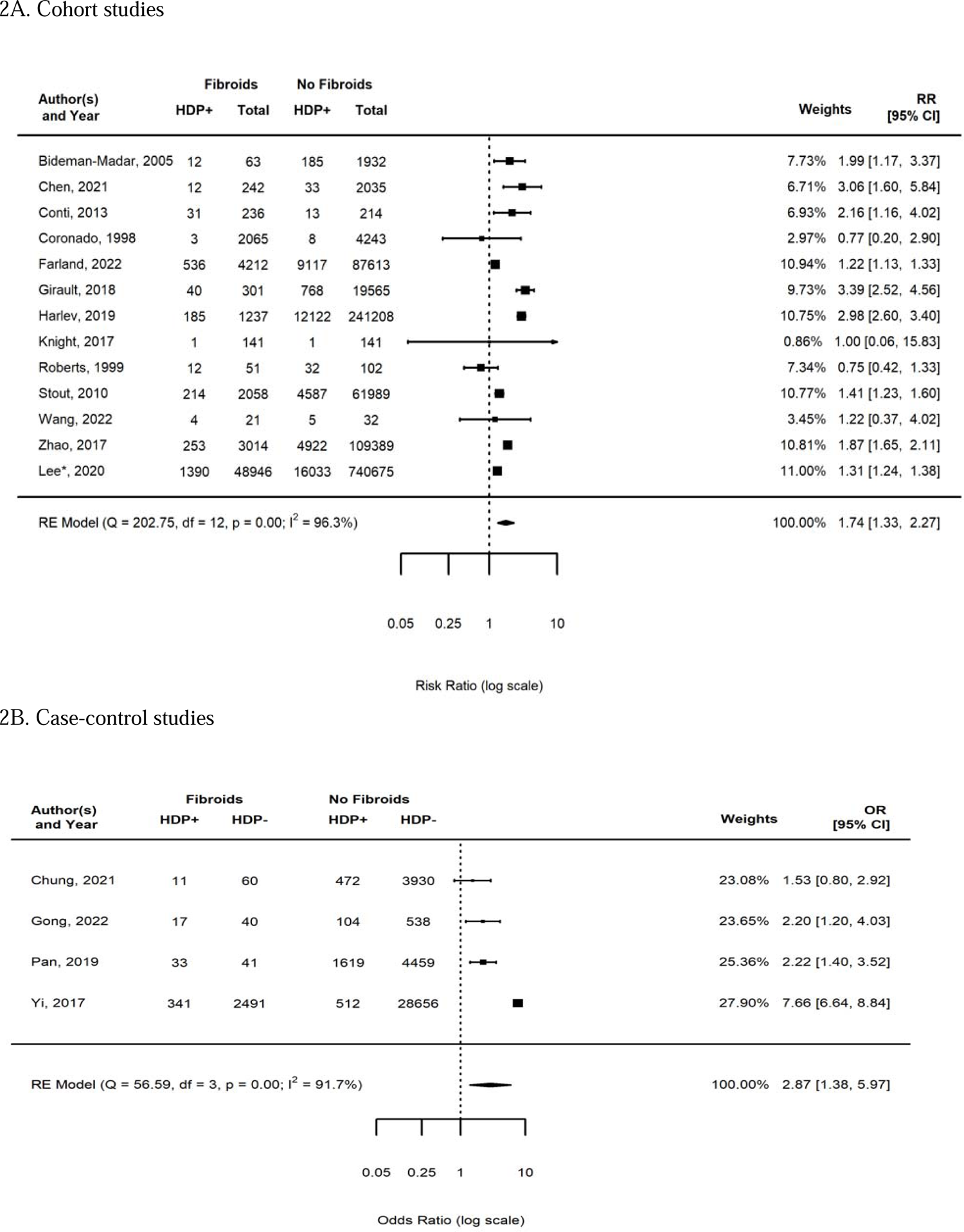
Forest Plots for the association between uterine fibroids and hypertensive disorders of pregnancy. 2A. Cohort studies

### Quality Assessment and Risk of Bias

A funnel plot and Egger’s test for retrospective studies did not show evidence of publication bias (P-value 0.5) (Figure 3). Excluding the intervention arm of the Lee et al (26) study did not change the publication bias among retrospective studies (Supplement 6). Regarding case-control studies, the funnel plot did show asymmetry indicating the possibility of publication bias; however, this was based on a very small number of studies (P-value <0.001) (Figure 3).

**Figure 3.**
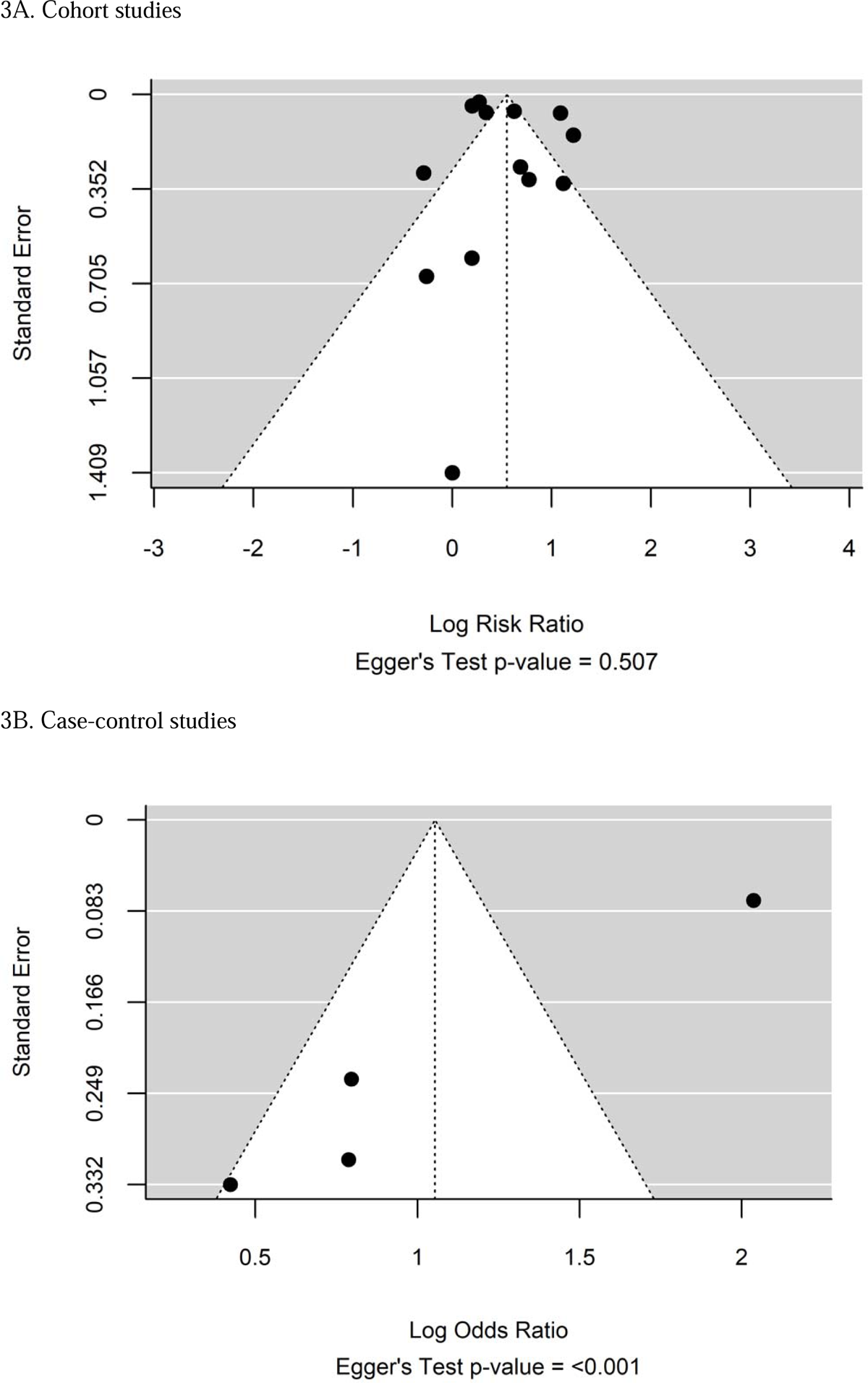
Funnel Plots for publication bias.

All included studies were high quality (NOS≥7) with the exception of three (17, 24, 27) (Table 2). We performed several additional sensitivity analyses including with the low-quality studies excluded and with and without the Lee intervention arm (26) and these did not change the direction or magnitudes of our estimates. Lastly, we performed sensitivity analyses with each study individually removed to see if perhaps one study was the primary driver of the association and our relative risks for cohort studies ranged from RR 1.54-1.81, and odds ratios for case-control studies ranged from OR 2.21-4.25, demonstrating that the results of the pooled estimates were very robust and persisted in each of these scenarios.

## DISCUSSION

This systematic review and meta-analysis comprised 17 available studies including 1,374,395 participants of whom 64,968 were diagnosed with uterine fibroids. In both retrospective cohort and case-control studies, a persistent association between uterine fibroids and an increased risk of HDP was present – 74% higher risk in cohort studies, and odds of uterine fibroids were 2.87 times higher in women with HDP than without.

The association between uterine fibroids and chronic hypertension has been extensively documented in the literature (8, 28–30), though causality has not been established (9). Possible explanations include shared risk factors (28), smooth muscle injury through mechanical shear stress as a predisposing factor for uterine vasculature inducing myomatous proliferation (31), or the proinflammatory milieu induced by elevated blood pressure (32, 33). Intriguingly, one nested case-control study from a private health insurance claims database demonstrated that women on an angiotensin-converting enzyme inhibitor experienced a greater than 30% reduced odds of developing clinically symptomatic uterine fibroids compared to non-users (34). This was in contrast to the examination of thiazide diuretics to exclude the possibility that the mechanism was mediated through control of blood pressure itself – in that group, women had an increased, not decreased, risk of fibroid diagnosis, potentially implicating aberrations in angiotensin-converting enzyme inhibitor targets-including angiogenesis and cell proliferation-in women with uterine fibroids (10). Furthermore, 3-hydroxy-3-methylglutaryl coenzyme A (HMG-CoA) reductase inhibitors used to treat hypercholesterolemia have been associated with a lower risk of uterine fibroids and fibroid-related symptoms in a retrospective cohort study, again suggesting aberrations in metabolic pathways associated with uterine fibroids (35). Therefore, uterine fibroids may now be perceived as a marker for hypertension, and some have suggested that women with fibroids may start to be screened for elevated blood pressure and vice versa (36).

Studies examining the association between uterine fibroids and HDP, synthesized in this report, are scarcer. Preeclampsia is a leading cause of maternal death and a major contributor to maternal and perinatal morbidity therefore identification of possible risk factors is critical. Hypotheses for the impact of uterine fibroids on the risk of HDP include disruption of trophoblastic invasion of spiral arteries caused by the expanding fibroid (21). However, the links between uterine fibroids and chronic hypertension above also suggest that there may be more than just a mechanical effect and that systemic vascular dysfunction may play a role. Given the strength of the association found in this meta-analysis, including its persistence in all sub-analyses, further study into the mechanism of this association is warranted.

### Strength and Limitations

This is the first meta-analysis, to our knowledge, that evaluated the association between uterine fibroids and hypertensive disorders in pregnancy. This report includes a large number of participants globally and all but 3 studies received high-quality scores using the NOS criteria. We performed meta-regressions to test whether the association between uterine fibroids and HDP was modified by age or BMI. Multiple sensitivity analyses were performed to ensure the robustness of our conclusion.

This study has several limitations. First, many of the included studies were European and Asian populations. Uterine fibroids pose a significant health burden that disproportionally effects Black women, and that group may have different risk profiles (37). We intended to perform meta regressions including race, but this was not possible due to the small number of studies that included information about this covariate.

Importantly, there was significant heterogeneity in the definition of hypertensive disorders of pregnancy (Supplement 1), and the way in which the uterine fibroid diagnoses were established (Table 1). We utilized a random-effects model in anticipation of the anticipated heterogeneity of the included studies.

Secondly, all the studies included in this meta-analysis were retrospective in nature; while we included adjustments for measured covariates in meta-regressions, there are limitations to retrospective studies that include unmeasured confounding. It is also important to note that in meta-analyses of aggregate data, associations between average patient characteristics and the pooled treatment effect do not necessarily reflect true associations between the individual patient-level characteristics and treatment effect, also known as ecological fallacy. Thus, the results of this study must be used with caution when applied to any individual.

Finally, while this work is meant to be hypothesis-generating, the association is not causation and further studies are needed to explore the relationship between uterine fibroids and HDP. Future studies could include investigating the impact of increased surveillance and preventative strategies for HDP in this population including acetylsalicylic acid.

## CONCLUSION

Uterine fibroids were associated with an increased risk of hypertensive disorders of pregnancy when all available literature was synthesized, including when shared risk factors are examined in meta-regression analyses. Investigations into the mechanisms of this association are needed as this finding potentially has implications for risk stratification and monitoring for hypertensive disorders of pregnancy in this population.

## Supporting information

Supplement 1 Search Strategy

Supplement 2 Definition of HDP

Supplement 4B Meta Reg Plot by BMI

Supplement 5A Meta Reg Plot Age no Lee

Supplement 5B Meta Reg Plot BMI no Lee

Supplement 6 Funnel Plot no Lee

Supplement 3 Forest plot no Lee

Supplement 4A Meta Reg Plot Age

## Data Availability

All data produced in the present study are available upon reasonable request to the authors.

