## Supplement 1 Search Strategy for "Uterine Fibroids and Hypertensive Disorders in Pregnancy: A Systematic Review and Meta-Analysis"

All searches were conducted through April 21, 2023 and limited to publications in English.

**Cochrane Library (Wiley) – 57 records**

([mh "Leiomyoma"] OR (fibroid OR fibroids OR fibroma OR fibromas OR fibromyoma OR fibromyomas OR leiomyoma OR leiomyomas OR leiomyomata OR leiomyomatas OR leiomyomatosis OR leiomyomatoses OR myoma OR myomas OR myomatosis):ti,ab,kw) AND ([mh "hypertension, pregnancy induced"] OR (([mh "Pregnancy"] OR [mh "Pregnancy Outcome"] OR (gestational OR obstetric OR pregnant OR pregnancy OR pregnancies OR maternal):ti,ab,kw) AND ([mh "Hypertension"] OR ("blood pressure" OR hypertension OR hypertensions OR hypertensive OR toxaemia OR toxaemias OR toxemia OR toxemias OR toxicosis):ti,ab,kw)) OR ("chronic hypertension" OR eclampsia OR eclampsias OR eclamptic OR eclamptogenic OR "edema proteinuria hypertension gestosis" OR "EPH complex" OR "EPH gestosis" OR "EPH syndrome" OR "EPH toxemia" OR gestosis OR "HELLP syndrome" OR "hemolysis elevated liver enzymes low platelet" OR "pre eclampsia" OR preeclampsia OR "pre eclamptic" OR preeclamptic OR "superimposed hypertension"):ti,ab,kw OR [mh "Pregnancy Complications"] OR ("adverse obstetric outcome*" OR "adverse pregnancy outcome*" OR "adverse outcome during pregnanc*" OR "adverse outcomes during pregnanc*" OR "poor obstetric outcome*" OR "poor pregnancy outcome*" OR "poor outcome during pregnanc*" OR "poor outcomes during pregnanc*" OR "complications during pregnanc*" OR "pregnancy complication*" OR "obstetric complication*"):ti,ab,kw OR [mh "Premature Birth"] OR ("premature birth*" OR "preterm delivery" OR "preterm deliveries"):ti,ab,kw)

**Embase (Elsevier on embase.com) – 1,391 records**

('leiomyoma'/exp OR fibroid:ti,ab,kw OR fibroids:ti,ab,kw OR fibroma:ti,ab,kw OR fibromas:ti,ab,kw OR fibromyoma:ti,ab,kw OR fibromyomas:ti,ab,kw OR leiomyoma:ti,ab,kw OR leiomyomas:ti,ab,kw OR leiomyomata:ti,ab,kw OR leiomyomatas:ti,ab,kw OR leiomyomatosis:ti,ab,kw OR leiomyomatoses:ti,ab,kw OR myoma:ti,ab,kw OR myomas:ti,ab,kw OR myomatosis:ti,ab,kw) AND ('maternal hypertension'/exp OR (('pregnancy'/exp OR 'pregnancy outcome'/de) AND 'hypertension'/exp) OR (((chronic OR gestational OR obstetric OR pregnant OR pregnancy OR pregnancies OR maternal OR superimposed) NEAR/3 ('blood pressure' OR hypertension OR hypertensions OR hypertensive OR toxaemia OR toxaemias OR toxemia OR toxemias OR toxicosis)):ti,ab,kw) OR eclampsia:ti,ab,kw OR eclampsias:ti,ab,kw OR eclamptic:ti,ab,kw OR eclamptogenic:ti,ab,kw OR gestosis:ti,ab,kw OR 'pre eclampsia':ti,ab,kw OR preeclampsia:ti,ab,kw OR 'pre eclamptic':ti,ab,kw OR preeclamptic:ti,ab,kw OR ((eph NEAR/3 (complex OR syndrome OR toxemia)):ti,ab,kw) OR 'hellp syndrome':ti,ab,kw OR 'hemolysis elevated liver enzymes low platelet':ti,ab,kw OR 'pregnancy complication'/exp OR (((adverse OR negative OR poor) NEAR/3 (gestational OR obstetric OR pregnancy OR pregnancies OR pregnant) NEAR/3 outcome*):ti,ab,kw) OR (((gestational OR obstetric OR pregnancy OR pregnancies OR pregnant) NEAR/3 complication*):ti,ab,kw) OR 'prematurity'/de OR (((premature OR preterm) NEAR/3 (birth* OR delivery OR deliveries)):ti,ab,kw)) AND [english]/lim

**PubMed – 1,673 records**

("Leiomyoma"[MeSH Terms] OR "fibroid"[Text Word] OR "fibroids"[Text Word] OR "fibroma"[Text Word] OR "fibromas"[Text Word] OR "fibromyoma"[Text Word] OR "fibromyomas"[Text Word] OR "Leiomyoma"[Text Word] OR "leiomyomas"[Text Word] OR "leiomyomata"[Text Word] OR "leiomyomatas"[Text Word] OR "leiomyomatosis"[Text Word] OR "leiomyomatoses"[Text Word] OR "myoma"[Text Word] OR "myomas"[Text Word] OR "myomatosis"[Text Word]) AND ("hypertension, pregnancy induced"[MeSH Terms] OR (("Pregnancy"[MeSH Terms] OR "Pregnancy Outcome"[Mesh] OR "gestational"[Text Word] OR "obstetric"[Text Word] OR "pregnant"[Text Word] OR "pregnancy"[Text Word] OR "pregnancies"[Text Word] OR "maternal"[Text Word]) AND ("Hypertension"[MeSH Terms] OR "blood pressure"[Text Word] OR "hypertension"[Text Word] OR "hypertensions"[Text Word] OR "hypertensive"[Text Word] OR "toxaemia"[Text Word] OR "toxaemias"[Text Word] OR "toxemia"[Text Word] OR "toxemias"[Text Word] OR "toxicosis"[Text Word])) OR "chronic hypertension"[Text Word] OR "eclampsia"[Text Word] OR "eclampsias"[Text Word] OR "eclamptic"[Text Word] OR "eclamptogenic"[Text Word] OR "edema proteinuria hypertension gestosis"[Text Word] OR "EPH complex"[Text Word] OR "EPH gestosis"[Text Word] OR "EPH syndrome"[Text Word] OR "EPH toxemia"[Text Word] OR "gestosis"[Text Word] OR "HELLP syndrome"[Text Word] OR "hemolysis elevated liver enzymes low platelet"[Text Word] OR "pre eclampsia"[Text Word] OR "preeclampsia"[Text Word] OR "pre eclamptic"[Text Word] OR "preeclamptic"[Text Word] OR "superimposed hypertension"[Text Word] OR "Pregnancy Complications"[MeSH Terms] OR "adverse obstetric outcome*"[Text Word] OR "adverse pregnancy outcome*"[Text Word] OR "adverse outcome during pregnanc*"[Text Word] OR "adverse outcomes during pregnanc*"[Text Word] OR "poor obstetric outcome*"[Text Word] OR "poor pregnancy outcome*"[Text Word] OR "poor outcome during pregnanc*"[Text Word] OR "poor outcomes during pregnanc*"[Text Word] OR "complications during pregnanc*"[Text Word] OR "pregnancy complication*"[Text Word] OR "obstetric complication*"[Text Word] OR "Premature Birth"[MeSH Terms] OR "premature birth*"[Text Word] OR "preterm delivery"[Text Word] OR "preterm deliveries"[Text Word]) AND (english[Filter])

**MEDLINE (Ovid) – 1,196 records**

| **#** | **Query** Ovid MEDLINE(R) and Epub Ahead of Print, In-Process, In-Data-Review & Other Non-Indexed Citations and Daily |
| --- | --- |
| 1 | Leiomyoma/ |
| 2 | (fibroid or fibroids or fibroma or fibromas or fibromyoma or fibromyomas or leiomyoma or leiomyomas or leiomyomata or leiomyomatas or leiomyomatosis or leiomyomatoses or myoma or myomas or myomatosis).mp.   [mp=title, abstract, original title, name of substance word, subject heading word, floating sub-heading word, keyword heading word, organism supplementary concept word, protocol supplementary concept word, rare disease supplementary concept word, unique identifier, synonyms] |
| 3 | 1 or 2 |
| 4 | Hypertension, Pregnancy-Induced/ |
| 5 | Pregnancy/ or Pregnancy Outcome/ |
| 6 | Hypertension/ |
| 7 | 5 and 6 |
| 8 | ((chronic or gestational or obstetric or pregnant or pregnancy or pregnancies or maternal or superimposed) adj3 (blood pressure or hypertension or hypertensions or hypertensive or toxaemia or toxaemias or toxemia or toxemias or toxicosis)).mp. |
| 9 | (eclampsia or eclampsias or eclamptic or eclamptogenic or gestosis or "pre eclampsia" or preeclampsia or "pre eclamptic" or preeclamptic).mp. |
| 10 | ((EPH adj2 (complex or syndrome or toxemia)) or HELLP syndrome or hemolysis elevated liver enzymes low platelet).mp. |
| 11 | Pregnancy Complications/ |
| 12 | ((adverse or negative or poor) adj3 (gestational or obstetric or pregnancy or pregnancies or pregnant) adj3 outcome*).mp. |
| 13 | ((gestational or obstetric or pregnancy or pregnancies or pregnant) adj3 complication*).mp. |
| 14 | Premature Birth/ |
| 15 | ((premature or preterm) adj3 (birth* or delivery or deliveries)).mp. |
| 16 | 4 or 7 or 8 or 9 or 10 or 11 or 12 or 13 or 14 or 15 |
| 17 | 3 and 16 |
| 18 | limit 17 to english language |

**Scopus (Elsevier) – 1,633 records**

TITLE-ABS-KEY ((fibroid OR fibroids OR fibroma OR fibromas OR fibromyoma OR fibromyomas OR leiomyoma OR leiomyomas OR leiomyomata OR leiomyomatas OR leiomyomatosis OR leiomyomatoses OR myoma OR myomas OR myomatosis) AND (((chronic OR gestational OR obstetric OR pregnant OR pregnancy OR pregnancies OR maternal OR superimposed) W/3 ("blood pressure" OR hypertension OR hypertensions OR hypertensive OR toxaemia OR toxaemias OR toxemia OR toxemias OR toxicosis)) OR eclampsia OR eclampsias OR eclamptic OR eclamptogenic OR gestosis OR "pre eclampsia" OR preeclampsia OR "pre eclamptic" OR preeclamptic OR (eph W/3 (complex OR syndrome OR toxemia)) OR "hellp syndrome" OR "hemolysis elevated liver enzymes low platelet" OR ((adverse OR negative OR poor) W/3 (gestational OR obstetric OR pregnancy OR pregnancies OR pregnant) W/3 outcome*) OR ((gestational OR obstetric OR pregnancy OR pregnancies OR pregnant) W/3 complication*) OR ((premature OR preterm) W/3 (birth* OR delivery OR deliveries)))) AND (LIMIT-TO (LANGUAGE,"English" ))

**Web of Science Core Collection (Clarivate) – 508 records**

Editions = A&HCI , BKCI-SSH , BKCI-S , CCR-EXPANDED , ESCI , IC , CPCI-SSH , CPCI-S , SCI-EXPANDED , SSCI

TS=((fibroid OR fibroids OR fibroma OR fibromas OR fibromyoma OR fibromyomas OR leiomyoma OR leiomyomas OR leiomyomata OR leiomyomatas OR leiomyomatosis OR leiomyomatoses OR myoma OR myomas OR myomatosis) AND (((chronic OR gestational OR obstetric OR pregnant OR pregnancy OR pregnancies OR maternal OR superimposed) NEAR/3 ("blood pressure" OR hypertension OR hypertensions OR hypertensive OR toxaemia OR toxaemias OR toxemia OR toxemias OR toxicosis)) OR eclampsia OR eclampsias OR eclamptic OR eclamptogenic OR gestosis OR "pre eclampsia" OR preeclampsia OR "pre eclamptic" OR preeclamptic OR (eph NEAR/3 (complex OR syndrome OR toxemia)) OR "hellp syndrome" OR "hemolysis elevated liver enzymes low platelet" OR ((adverse OR negative OR poor) NEAR/3 (gestational OR obstetric OR pregnancy OR pregnancies OR pregnant) NEAR/3 outcome*) OR ((gestational OR obstetric OR pregnancy OR pregnancies OR pregnant) NEAR/3 complication*) OR ((premature OR preterm) NEAR/3 (birth* OR delivery OR deliveries)))) and English (Languages)
