## Supplement 2 Definition of HDP for "Uterine Fibroids and Hypertensive Disorders in Pregnancy: A Systematic Review and Meta-Analysis"

Table 1. Definition of Hypertensive Disorders of Pregnancy in Each Study

| Study | Definition |
| --- | --- |
| Biderman-Madar | Pregnancy induced hypertension^a^ |
| Chen | Gestational Hypertension and Preeclampsia^b^ |
| Chung | Answer “YES” to the survey question^c^ |
| Conti | Gestational hypertension^d^ |
| Coronado | Preeclampsia^e^ |
| Gong | Preeclampsia^f^ |
| Farland | PIH or eclampsia or preeclampsia^g^ |
| Girault | Gestational Hypertension and Preeclampsia^h^ |
| Harlev | Hypertensive disorders^i^ |
| Knight | Gestational hypertention^i^ |
| Lee | Preeclampsia^i^ |
| Pan | Preeclampsia^i^ |
| Roberts | Preeclampsia^j^ |
| Stout | Preeclampsia^k^ |
| Wang | HDCP and Preeclampsia^i^ |
| Yi | Preeclampsia^l^ |
| Zhao | Gestational hypertension, Preeclampsia, Eclampsia, Chronic hypertension^i^ |

SBP, systolic blood pressure, DBP, diastolic blood pressure , PIH, pregnancy induced hypertension, HDP, hypertensive disorders of pregnancy, HDCP, hypertensive disorder complicating pregnancy

a: Pregnancy induced hypertension by Davey and MacGillivray Classification

b: Gestational hypertension was defined as SBP at least 140 mmHg and/or DBP at least 90 mmHg without proteinuria, which develops after 20 weeks of gestation. Preeclampsia was defined as SBP at least 140 mmHg and/or DBP at least 90 mmHg after 20 weeks of gestation and proteinuria (defined as >300 mg of protein in a 24-h urine specimen or >1+ in two random urine samples collected at least 4 h apart)

c: Survey question: “Were you diagnosed or treated for hypertension (high blood pressure) during pregnancy?”

d: Reported by patient in the questionnaire

e: Recorded as pregnancy complication on the birth certificate

f: Preeclampsia was defined according to the International Society for the Study of Hypertension in Pregnancy (ISSHP) recommendation as SBP ≥140 mmHg and/or DBP ≥90 mmHg after 20 weeks gestation, combined with at least one of the following new-onset conditions: (1) positive proteinuria (protein–to–creatinine ratio of ≥30 mg/mmol or urinary dipstick ≥2+); (2) other maternal organ dysfunction (serum creatinine ≥90 μmol/L, alanine aminotransferase or aspartate aminotransferase >40 IU/L); (3) neurological complications (eclampsia, altered mental state, blindness, stroke, clonus, severe headaches, or persistent visual scotomata); (4) hematological complications (thrombocytopenia-platelet count <150,000/μL, disseminated intravascular coagulation, and hemolysis); and (5) uteroplacental dysfunction (fetal growth restriction, abnormal umbilical artery Doppler waveform analysis, or stillbirth)

g: Information on hypertension during pregnancy, was identified in Massachusetts Pregnancy to Early Life (PELL) from either the birth certificate or the hospital discharge delivery record and pre delivery (within 280 days before delivery) ICD-9 and ICD-10 codes (642.3-642.6,642.9, O11, and O13-O16 for pregnancy-related hypertension). PIH (pregnancy induced hypertension)

h: Collected from electronic medical records

i: Not specified in the article

j: Preeclampsia was defined as 2 blood pressures taken at least 6 hours apart that were equal to or exceeded 140 mm Hg systolic and/or 90 mm Hg diastolic or a rise in systolic blood pressure in a patient with chronic hypertension.

k: Defined by the American College of Obstetricians and Gynecologists criteria (citation)

l: Based on the medical records, each healthy pregnant woman who was diagnosed pre-eclamptic before labor pain was defined as case, and who were not diagnosed pre-eclamptic was defined as control. The pregnant women who were not diagnosed hypertensive before 2nd trimester, but their blood pressure was higher than 140/90 mmHg in last trimester with urine albumin level of more than 300 mg were in preeclampsia cases group, those who had no higher blood pressure and urine albumin level were considered in control group.
