## Supplementary figures and images for "Uterine Fibroids and Hypertensive Disorders in Pregnancy: A Systematic Review and Meta-Analysis"

### Supplement 3 Forest plot no Lee

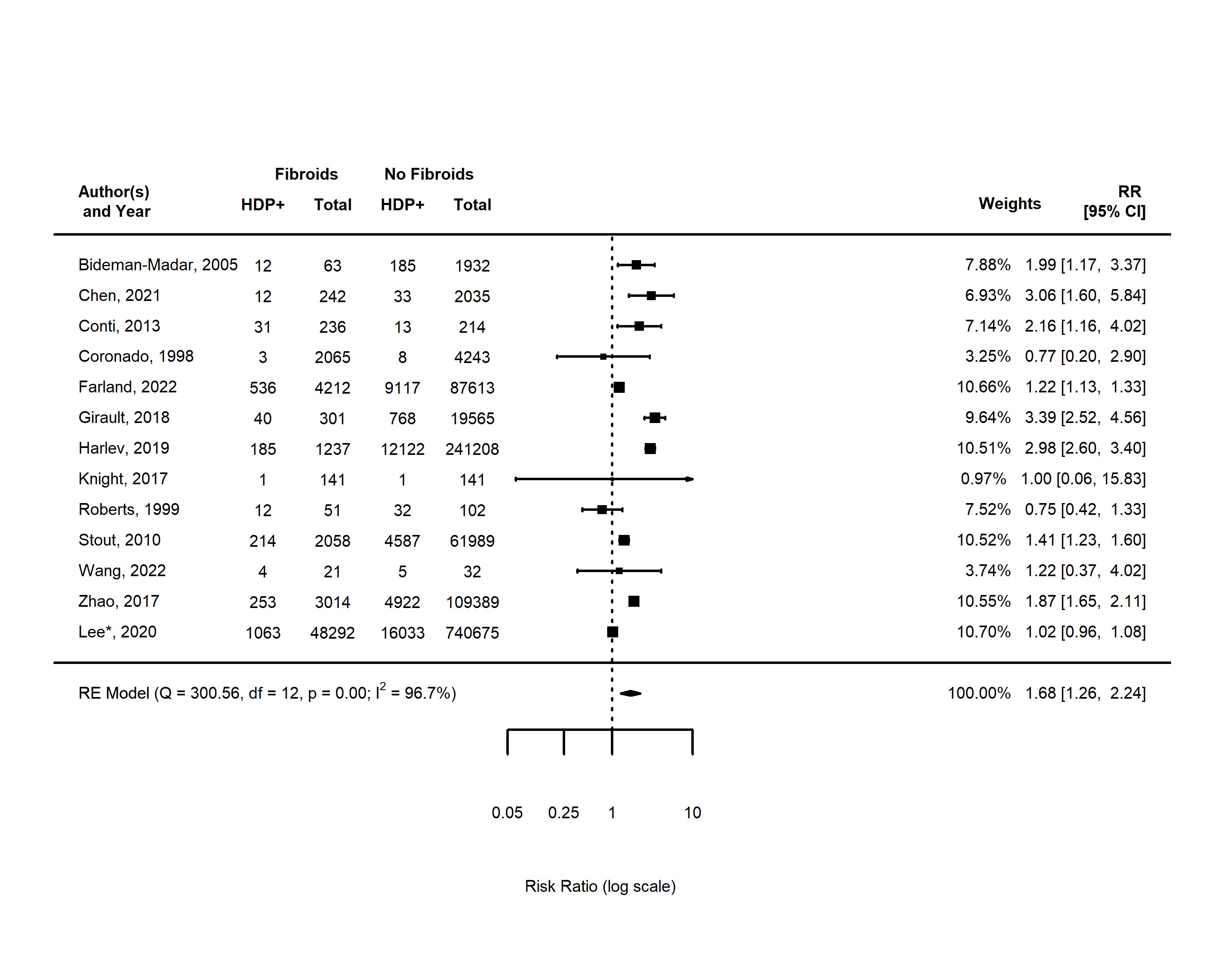

### Supplement 4B Meta Reg Plot by BMI

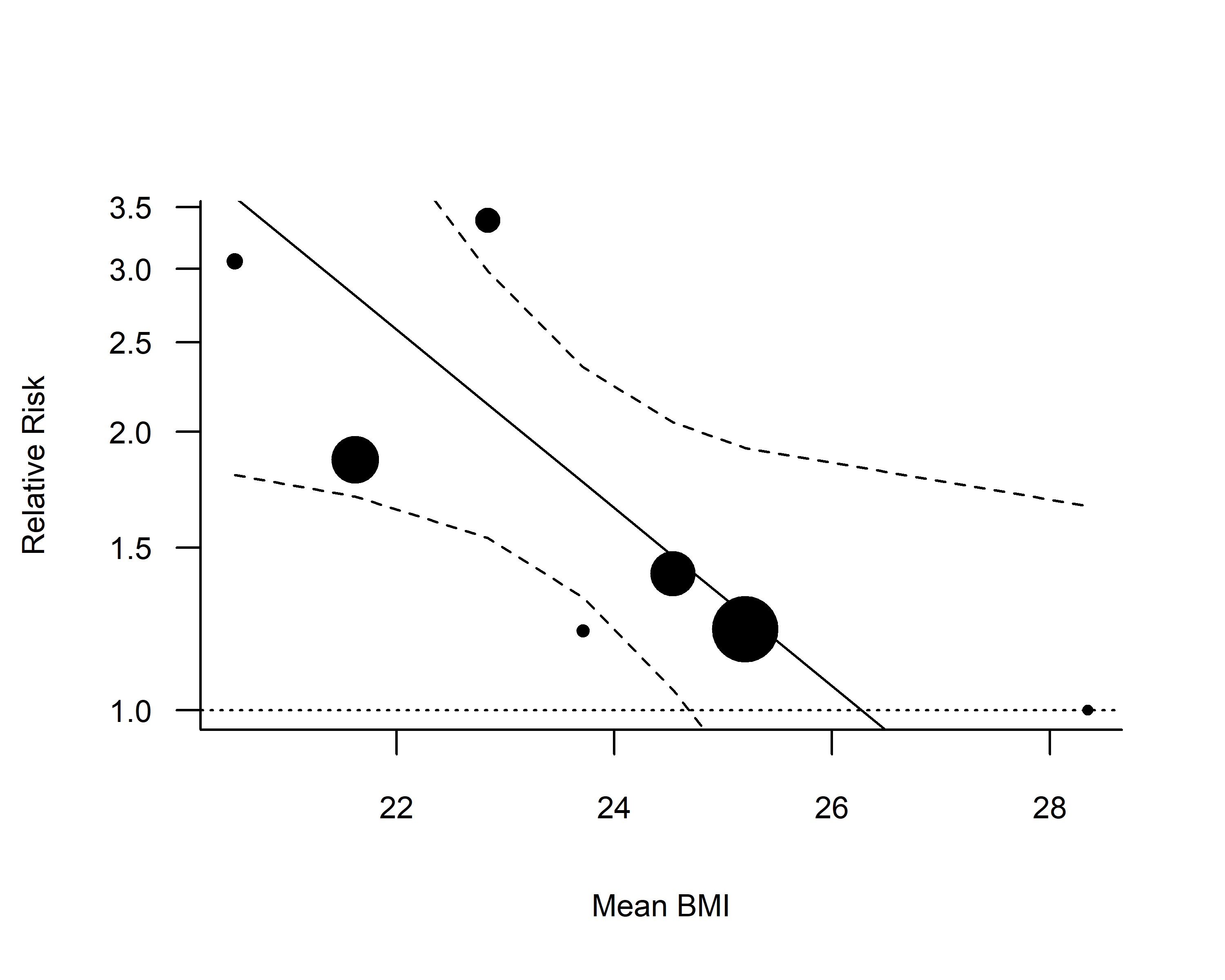

### Supplement 5A Meta Reg Plot Age no Lee

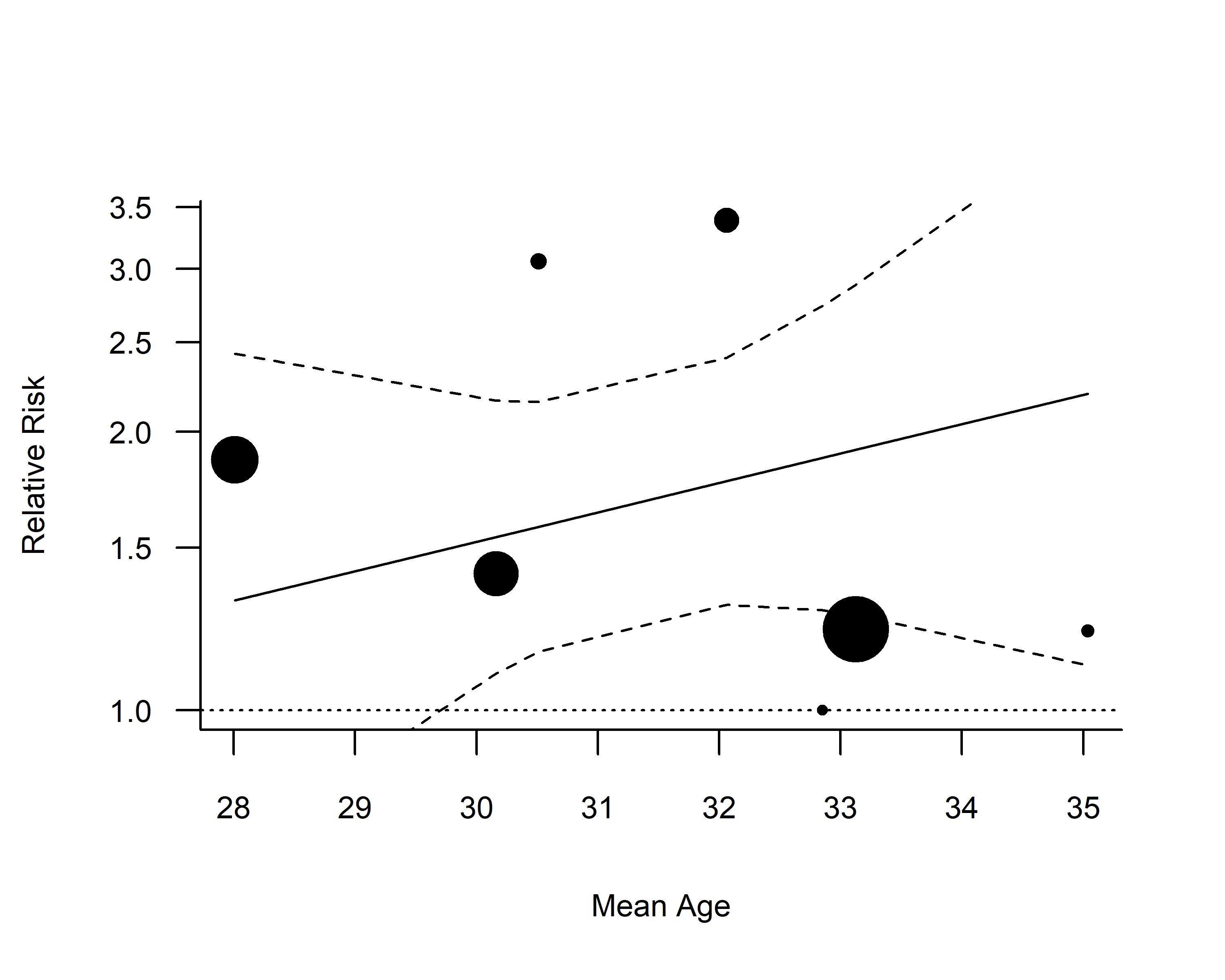

### Supplement 6 Funnel Plot no Lee

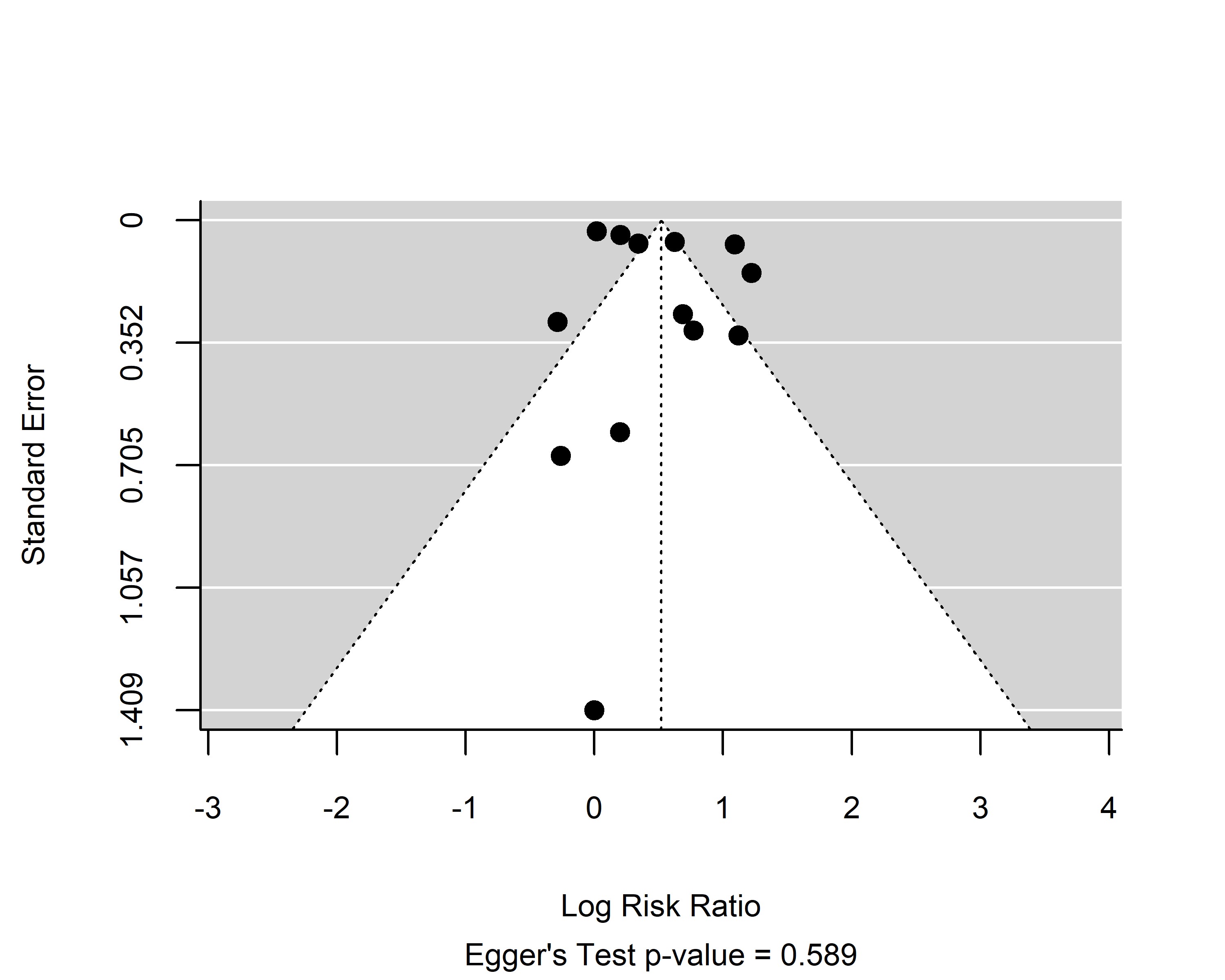
